# Antimicrobial concentrations in the cornea and aqueous humour: a meta-analysis

**DOI:** 10.1101/2025.05.16.25327782

**Authors:** Keri McLean, Grace Dawson, Daniel Foulkes, Rose Herbert, Petya Popova, Danielle Bernard-Deshong, Victoria Massie, Alfredo Borgia, Matteo Airaldi, Tobi Somerville, Gabriela Czanner, Timothy Neal, Stephen Tuft, Stephen B Kaye

**Affiliations:** Liverpool University Hospitals Foundation Trust, Liverpool, UK; Department of Eye and Vision Science, Institute of Life Course and Medical Science, University of Liverpool, Liverpool, UK; School of Medicine, University of Sunderland, Sunderland, UK; School of Medicine, University of Liverpool, Liverpool, UK; UCL Institute of Ophthalmology, London, UK; School of Electronics and Computer Science, Faculty of Medicine, University of Southampton, Southampton, UK

**Keywords:** Cornea, keratitis, antimicrobial, concentration

## Abstract

**Aim:** To interpret the likely clinical susceptibility of isolates from microbial keratitis we performed a meta-analysis of published data that measured the concentrations of, topically applied, antimicrobials in the cornea or aqueous humour. We then correlated these values with the *in vitro* minimum inhibitory concentration (MIC).

**Methods and Analysis:** We searched PubMed to identify studies reporting aqueous and/or corneal concentrations of 53 topically applied ocular antimicrobials, spanning the following classes: beta-lactams, glycopeptides, aminoglycosides, chloramphenicol, lincosamides, macrolides, oxazolidinones, steroidal antimicrobials, tetracyclines, diaminopyrimidines, sulfonamides, lipopeptides and polymyxins. For each class, two clinicians independently screened the abstracts. For articles that met the inclusion criteria, data were extracted on participant species, antimicrobial concentration, dosing regimens, epithelial status, and measurement methods. Concentrations were standardised to mg/L. First quartile concentrations (EQ1) were extrapolated from the mean and standard deviation or calculated from the contained data where available. The data were tabulated to generate the EQ1 concentrations for the aqueous and cornea of each antimicrobial, stratified by applied concentration and dosing regimen.

**Results:** We screened 7247 publications. Eighty-one publications were included in the meta-analysis, comprising data on the aqueous and/or corneal concentrations of twenty-eight antimicrobials. Bioassay was the most frequently used method for quantifying antimicrobial concentrations (25 studies), followed by liquid chromatography and fluorescence assays (18 studies each), mass spectrometry (12 studies), and radioactivity and colorimetric assays (three studies each).

**Conclusion:** We provide a practical resource for clinicians to assess whether the expected EQ1 of an antimicrobial in the cornea is above the *in vitro* MIC of the pathogen. This reduces reliance on systemic break-point concentrations enabling evidence-based antimicrobial treatment decisions for microbial keratitis.

**KEY MESSAGES:** *What is already known on the topic:* Microbial keratitis (MK) is a major cause of preventable blindness worldwide. The susceptibility of an isolate is based on systemic breakpoint criteria that may not reflect corneal or aqueous concentrations following topical application.

*What this study adds:* We provide a comprehensive and standardised resource of corneal and aqueous antimicrobial concentrations following topical application. This enables treatment decisions based on the minimal inhibitory concentrations (MIC) of the isolate and the expected tissue concentration of the antimicrobial for evidence-based management of MK.

*How this study might affect research, practice or policy:* This study provides a practical resource linking *in vitro* antimicrobial MIC values to anticipated ocular drug concentrations, enabling more precise treatment of microbial keratitis, supporting research and the development of clinical guidelines.

## INTRODUCTION

Microbial keratitis (MK) is a preventable cause of blindness resulting from infection of the cornea by bacteria, fungi, protozoa or viruses. Global prevalence estimates vary widely from 4.5 – 37.7 cases per 100 000 in high-income countries compared to 113 – 362 per 100 000 in low- and middle-income countries.^(1, 2)^ MK is associated with painful corneal ulceration and inflammation, scarring, perforation and loss of vision.^(3)^ Prompt diagnosis and treatment are critical to preserve sight. Samples are collected for identification of the causative microorganism with antimicrobial susceptibility tests to guide therapy. Since these processes typically take 24–48 hours or longer, empirical antimicrobial therapy is commenced at presentation and then adjusted according to the results of the investigations and the clinical response.

Microbiology reports provide clinicians with genus and/or species of the microorganism and its susceptibility to antimicrobials based on systemic breakpoint concentrations. Importantly, reports generally omit the minimal inhibitory concentrations (MIC) of the antimicrobial against the pathogen. Instead, susceptibility is classified as susceptible, intermediate (susceptible at increased concentration) or resistant, using standardised breakpoint guidelines, such as those from the Clinical and Laboratory Standards Institute (CLSI) or European Committee for Antimicrobial Susceptibility Testing (EUCAST).(^4, 5^) These guidelines are based on the anticipated response of the bacteria against the antimicrobial concentrations that can be achieved in the serum following systemic administration, but do not reflect the concentrations achieved in the cornea or aqueous humour after topical administration.(^3, 6^) As a result, systemic breakpoints seldom apply to MK.(^6, 7^) Because there is an association between the clinical outcome and the MIC of the topical antimicrobial used to treat the MK,^(8,9)^ comparing the MIC of antimicrobials against the causative organism and the achievable corneal concentration following topical application may provide more relevant guide to treatment.

To utilise MIC values effectively, clinicians need to know the achieved concentrations of topically applied antimicrobials in the cornea and aqueous humour, as well as the factors which can affect these levels, such as an intact or abraded corneal epithelium. Data from animal and human studies show that the concentration can vary significantly with different dosing regimens, measurement methods, and reporting units. This has led to uncertainty as to the expected antimicrobial concentration in the cornea and aqueous following topical application. In addition, the MIC is typically expressed in mg/L (equivalent to μg/ml), whereas corneal concentrations are often reported as mg/g of cornea. Although they can be converted, standardising the reporting of these units would provide more easily interpretable and actionable data for clinicians.

In this study we performed a meta-analysis and collated the published data on the corneal and aqueous concentrations of antimicrobials measured following topical application. For each antimicrobial, we provide an overview of their function and their corneal and aqueous concentrations in mg/L. We have also provided the first-quartile concentration (EQ1)^(1)^ as the expected concentration is very unlikely to be less than the EQ1. Hence, if the MIC of the pathogen is less than the EQ1 it would indicate that the isolate was susceptible to the respective antimicrobial. ^(1)^

## METHODS

A PubMed search, was completed on the November 8, 2024 using the following search terms, (((keratitis[Title/Abstract]) OR (cornea[Title/Abstract])) OR (ocular[Title/Abstract])) AND (*insert relevant antimicrobial*[Title/Abstract]). The antimicrobials were: aberkacin, amikacin, azithromycin, aztreonam, carbenicillin, cefazolin, cefaloridine, cefepime, cefmenoxime, cefotaxime, ceftazidime, ceftriaxone, cefuroxime, chloramphenicol, clarithromycin, clindamycin, colistin (polymyxin E), daptomycin, doripenem, doxycycline, ertapenem, erythromycin, fusidic acid, gentamicin/gentamycin, imipenem, isepamycin, kanamycin, linezolid, lincomycin, meropenem, methicillin, metronidazole, minocycline, neomycin, netilmycin, oxacillin, penicillin, piperacillin, polymyxin B, rifampicin, soframycin, streptomycin, sulfacetamide, sulfamethoxazole, sulfisoxazole, teicoplanin, teixobactin, tetracycline, tigecycline, ticarcillin, tobramycin, trimethoprim/sulphamethoxizole, and vancomycin. Abstracts were screened by two clinicians to select studies reporting antimicrobial concentrations after topical application in any species. We excluded fluoroquinolones as this data has been reported previously.^(1)^

The extracted data included the number and species of participants, concentration of the topically applied antimicrobial, dosing regimen, epithelial status (intact or abraded), measurement methods, concentrations and standard deviation (SD). Concentrations were standardised to mg/L (equivalent to μg/ml), the preferred unit for reporting minimal inhibitory concentrations (MICs). The percentage concentration weight per volume (w/v %) using a corneal density estimate of 1.087 kg/L^(10)^ and/ or volume per volume (v/v %) are also provided. Where possible, aqueous and corneal concentrations at one-hour post-instillation were recorded, or the concentration closest to this time point. Where only standard error (SE) was provided, SD was calculated in each study using the formula: 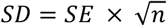.

Studies were grouped by species, antimicrobial and applied concentrations. For each paper, we calculated the extrapolated first quartile (EQ1) of the concentration. Where the actual data were not provided, the EQ1 was calculated assuming a normal distribution, using the formula: 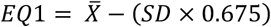.^(11)^ Weighted pooled SDs were derived using: 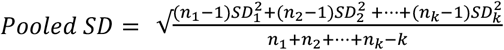, where n is the number of samples in each study and k is the total number of studies.^(11)^ The coefficient of variation (CoV) of the pooled data was calculated and expressed as a percentage using the following formula: 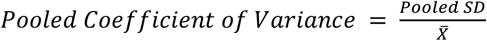. Analysis was conducted using Microsoft Office 365 Excel.

## RESULTS

Our search identified 7247 papers. Eighty-one publications were included in the meta-analysis, providing data on the corneal and aqueous concentrations of 28 antimicrobials (see PRISMA flow diagram).

**Figure 1:**
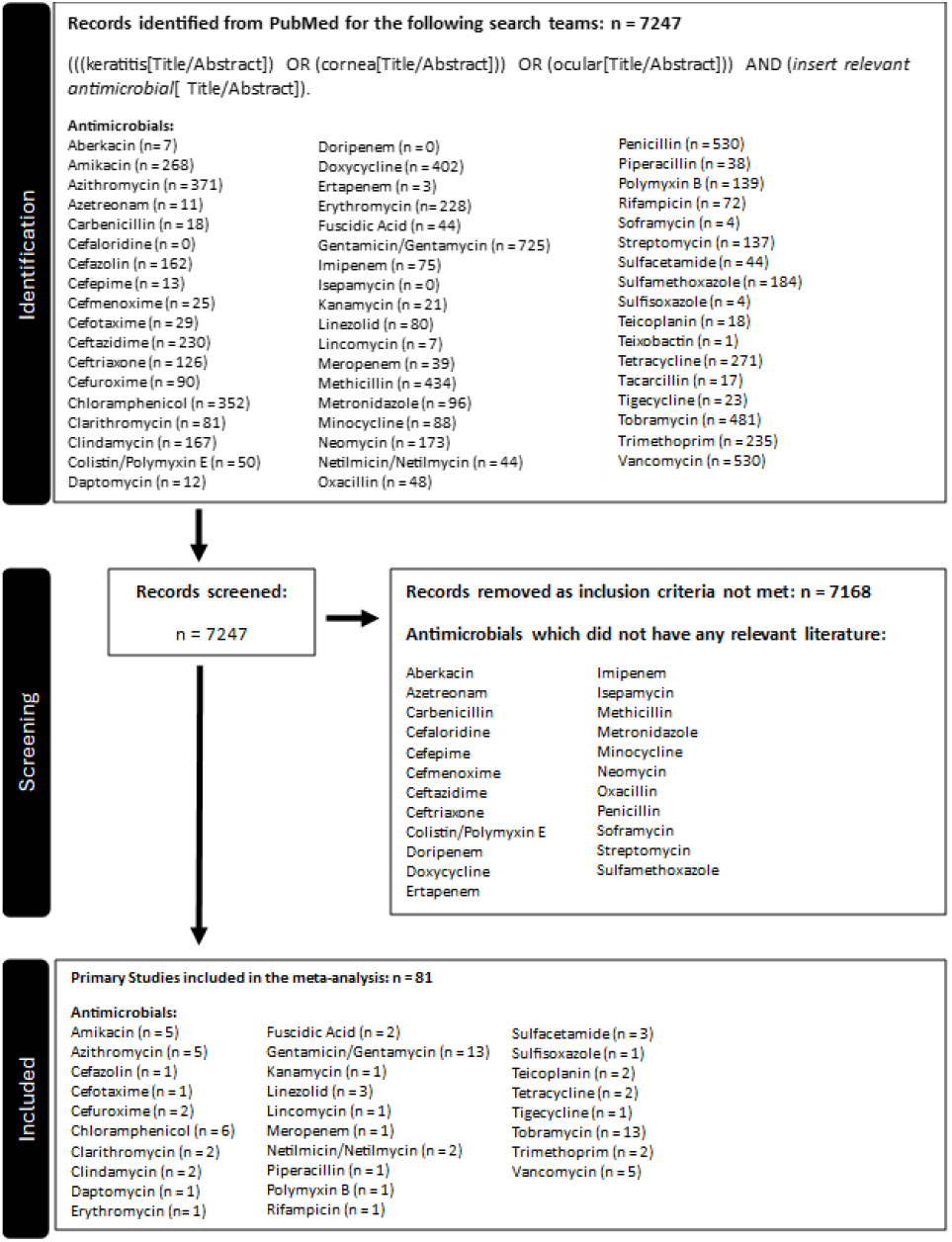
PRISMA flow diagram for meta-analysis of corneal and aqueous concentrations of antimicrobial following topical application.

A summary of the concentrations and method of measurement e.g., bioassay, chromatography, radioimmunoassay for each antimicrobial is provided in Supplementary Material 1 – Appendix 1. The mechanism of action for each antimicrobial class and drug have been complied in Supplementary Material 1 – Appendix 2. The extracted data and analysis, including references to the included papers, are compiled for each antimicrobial in Supplementary Material 2 (a .xlsx file).

The 81 publications included 33 studies conducted in humans, 47 in rabbits, and one in mixed-breed dogs. Bioassays were the most frequently used method for quantifying antimicrobial concentrations (30.9%, 25/81), followed by liquid chromatography (LC) and fluorescence assays (22.2%, 18/81 each). Mass spectrometry (MS) was utilised in 12 studies (14.8%, 12/81), while radioactivity and colorimetric assays were the least common, with three studies each (3.7%, 3/81 each). Dosing regimens varied widely from to once a day to four times a day (QDS).

Table 1 provides a summary of the corneal and aqueous EQ1 concentrations of each antimicrobial at one hour following application, stratified by the concentration of topically applied drug. EQ1 values are presented in mg/L, with the CoV for the pooled data expressed as a percentage. For this summary table, data from human studies was provided when available, but when human data were not available animal data has been included. This table is intended as a practical resource for clinicians to assess whether the antimicrobial’s EQ1 is above the MIC of the pathogen. Further details on its application can be found in the Discussion.

**Table 1:**
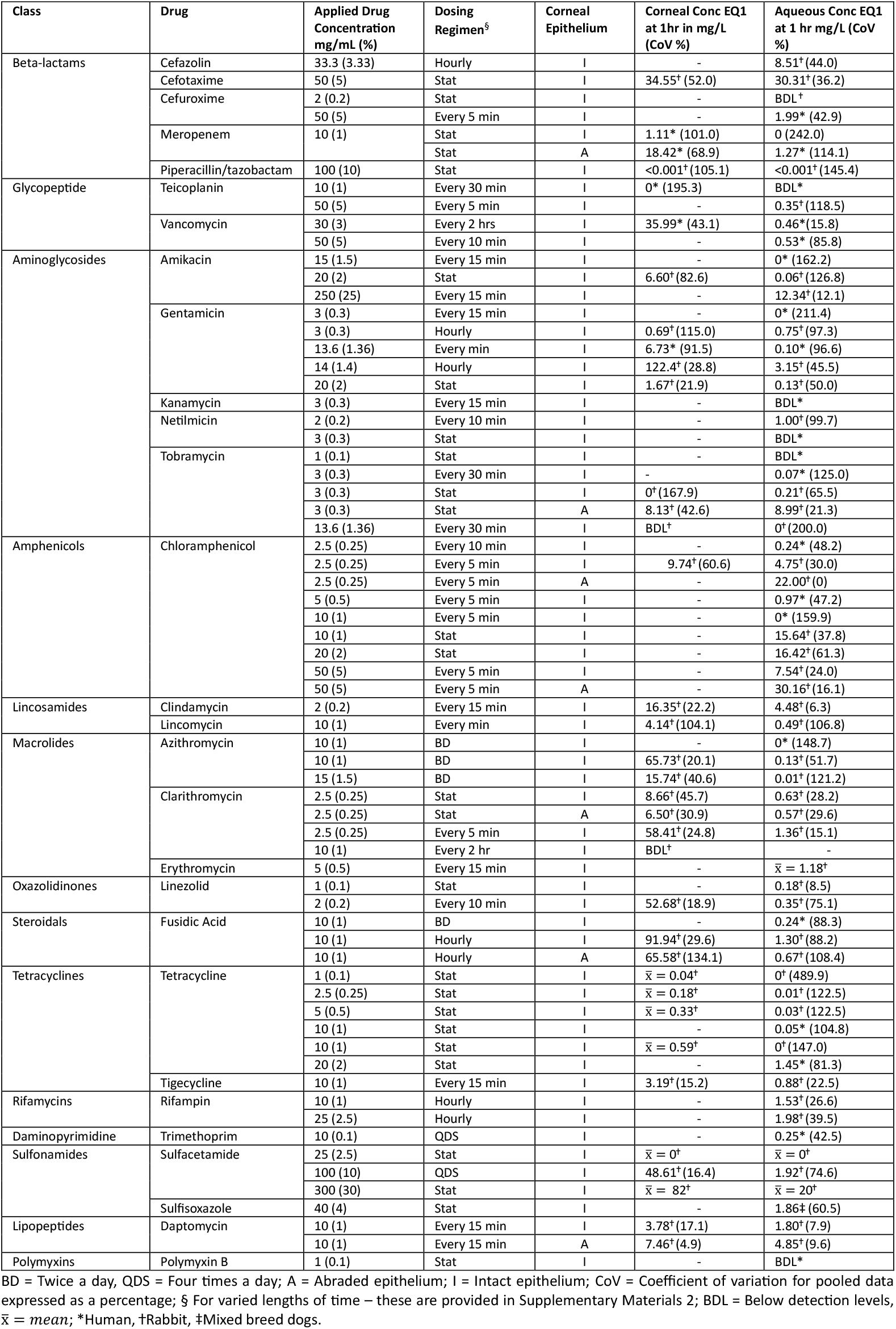
Summary Table of Corneal and Aqueous Extrapolated First Quartile (EQ1) Concentrations for Topically Applied Antimicrobials.

**Table 2:** Comparison of the published EUCAST resistance MIC breakpoints of *Pseudomonas aeruginosa, Staphylococcus aureus and Streptococcus pneumonia* with the corneal EQ1 of various antimicrobials.

|  | EUCAST MIC Resistance Breakpoint (mg/L) |  |  | Corneal EQ1 (mg/L) |
| --- | --- | --- | --- | --- |
|  | *Pseudomonas aeruginosa* | *Staphylococcus aureus* | *Streptococcus pneumonia* |  |
| Ciprofloxacin 3 mg/mL | 0.5 | 2 | 4 | 2.89* |
| Ofloxacin 3 mg/mL | 2 | 1 | 4 | 5.76* |
| Meropenem 10 mg/mL | 8 | NA | 2 | 18.42 (Abraded) |
| Amikacin 20 mg/mL | 16 | 16 | NA | 6.60 |
| Gentamicin 3 mg/mL | 8 | 2 | NA | 0.69 |
| Tobramycin 3 mg/mL | 2 | 2 | NA | 8.13 (Abraded) |
| Chloramphenicol 2.5 mg/mL | NA | 16 | 8 | 9.74 |
| Linezolid 2 mg/mL | NA | 4 | 2 | 52.68 |
| Fusidic Acid 10 mg/mL | NA | 0.5 | NA | 65.58 (Abraded) |

NA – Not available; \* Previously published data – Reference 1.

Data on the current systemic breakpoint concentrations for each microorganism and antimicrobial (where available) can be accessed from EUCAST.^(5)^ To demonstrate the benefit of using corneal concentrations we have, where available, compared the systemic breakpoint data to the corneal concentrations (EQ1) for three main pathogens, *P. aeruginosa, S. aureus and S. pneumonia* for several antimicrobials. For example, *P. aeruginosa* has a published EUCAST resistance MIC of 0.5 mg/L to ciprofloxacin and 8.0 mg/L to meropenem. The corneal EQ1s are however, 2.89 mg/L for ciprofloxacin and 18.42 mg/L for meropenem (abraded). As the corneal EQ1s are substantially higher than the systemic breakpoints one would expect bacteria with an MIC below these levels to be susceptible with topical treatment. *Staphylococcus aureus* has a published EUCAST resistance MIC of 16 mg/L to both amikacin and chloramphenicol. The corneal EQ1s of these antimicrobials are lower than the systemic breakpoints, therefore one would expect the bacteria to be less susceptible with topical treatment.

## DISCUSSION

Current microbial susceptibility guidelines, such CLSI or EUCAST, rely on breakpoint thresholds derived from serum antimicrobial concentrations achieved after systemic administration. These systemic concentrations, however, do not accurately reflect the pharmacokinetics of antimicrobials administered topically to the eye, particularly in achieving therapeutic levels within the corneal and aqueous compartments. This misalignment highlights the need for clinically relevant data that allow measured MIC values to be compared with estimates of the ocular penetration of antimicrobials, thus enabling more precise and potentially more effective treatment of MK. We have addressed this knowledge gap by summarising the corneal and aqueous concentrations of various topically applied ocular antimicrobials, thus creating a practical resource for clinicians.

### Reasoning for Table 1 Methodology

The coefficient of variation (CoV) shown in Table 1 expresses the degree of variability in the pooled data as a percentage of the mean EQ1 concentration. It provides insight into the consistency of reported antimicrobial concentrations across the different studies. A lower CoV value indicated more uniform data, suggesting the concentration given may be more reliable. Higher CoV values reflect greater variability within the pooled studies. This may be due to differences in study design, species, dosing regimens and measurement methods.

Recognising the inherent variability of the published literature—stemming from differences in study design, dosing regimens, and measurement techniques—our methodology extrapolates first-quartile concentrations (EQ1) for each antimicrobial and species (human or animal). All data were standardised as mg/L, the unit commonly used to report clinical MIC results. This approach ensures a conservative estimate of ocular antimicrobial levels, offering a more reliable reference point for clinicians to compare against the MIC of pathogens causing MK rather than using the expected mean concentration particularly when the variance of measurements is high. For example, referring to the EQ1, it would be expected that 75% of concentrations are greater than the EQ1. If as an alternative one were to set a stricter threshold of 2 SD below the mean or to use the lower 95% confidence limit, although this would guarantee that at least 75% of concentrations are above that level (Chebyshev’s theorem) due to the high CoV this would give very low or close to zero cutoff concentrations.

A variety of methods were used to measure the antimicrobial concentrations in the cornea and aqueous. Bioassays were the most frequently used method for quantifying antimicrobial concentrations, employed in 25 studies, followed by liquid chromatography (LC) and fluorescence assays, each used in 18 studies. Mass spectrometry (MS) was utilised in 12 studies, while radioactivity and colorimetric assays were the least common. Each of these methods have their limitations, vary in sensitivity and specificity and can make comparisons difficult. We have summarised below the strengths and weakness of the assays used to enable the reader to evaluate the presented data (Supplementary Material 1 -Appendix 3).

### Clinical Application of Corneal and Aqueous Concentrations in Table 1

- The MIC of all isolates to the respective antimicrobial should be provided.
- **Antimicrobial selection:** If the MIC of an isolated pathogen exceeds the EQ1 concentration stated in Table 1 for a specific drug, the likelihood of therapeutic success diminishes. In such cases, clinicians should either increase the concentration and frequency of the antimicrobial, and/or consider alternative antimicrobials appropriate to the microorganism with superior corneal or aqueous penetration. If combination?treatment is used, consideration should be given to those antimicrobial combinations which are additive or synergistic.^(12)^
- **Infection location:** The anatomical site of infection—whether limited to the superficial cornea, deeper stromal layers, or extending into the anterior chamber—and intact epithelium should guide antimicrobial selection. For instance, removal of epithelium to aid penetration and choice of antimicrobial e.g., meropenem, with high corneal penetration post-abrasion (EQ1: 18.42 mg/L), may be particularly effective in treating deeper stromal infections.
- **Personalised therapy:** Directly comparing MIC values with EQ1 enables ophthalmologists to tailor antimicrobial regimens, ensuring that drug concentrations consistently exceed the pathogen’s MIC to maximize therapeutic efficacy.

### Limitations and future Directions

While EQ1 represents a valuable clinical tool, it is not without limitations:

- **Variability in experimental conditions may contribute to discrepancies in reported ocular drug concentration:** Differences in drug retention, dosing protocols, and measurement techniques can affect the measured concentrations. For example, inadequate washing of corneal tissue prior to analysis may lead to measurement of residual surface drug rather than drug which has penetrated the cornea. Additionally, in studies where corneal tissue was obtained from penetrating keratoplasty procedures, the clinical indication for surgery was not always reported. This is an important limitation, as the quality and biomechanics of corneal tissue can vary substantially between conditions such as keratoconus, pseudophakic bullous?keratopathy and microbial keratitis, potentially influencing drug permeation and retention outcomes.
- **Lack of raw data:** The absence of raw data in many studies limits the precision of EQ1 extrapolations.
- **Inter-species differences:** Variations in ocular pharmacokinetics between human and animal models can influence the generalisability of using corneal and aqueous concentrations.

To address these challenges, future research should prioritise:

- **Improved methodologies:** Developing more reliable and accurate methods of measuring drug concentrations in the cornea and aqueous would be highly beneficial.
- **Standardised reporting:** Implementing uniform methodologies for antimicrobial penetration studies and to provide the actual data to aid future meta-analyses. Moreover, standardising units to express antimicrobial concentration, preferably in mg/L, would enhance the clarity, comparability, and clinical applicability of study findings.
- **Comprehensive databases:** Developing and regularly updating a centralised database of EQ1 values for a wide range of antimicrobials by groups such as EUCAST will enhance accessibility and utility for clinicians.

## Conclusion

By providing a practical framework for interpreting MIC values alongside using the expected ocular drug concentrations, empowers clinicians to make informed and evidence-based decisions in the management of MK. Integrating this metric into clinical practice will help bridges the gap between laboratory data and real-world applications.

## Data Availability

All data produced in the present study are available upon reasonable request to the authors.

## ETHICAL APPROVAL

Not applicable.

## AUTHOR CONTRIBUTIONS

Study concept and design: KM and SBK. Data collection: KM, GD, RH, PP, DBD, VM, AB, MA, TS and SBK. Data analysis: KM, GD and SBK. Data interpretation: KM, GC, TN, ST and SBK. Manuscript drafting: KM, GD and DF. Critical revision of the manuscript: KM, GC, TN, ST and SBK. All authors approved the final manuscript.

